# Demographic and occupational differences in physical activity and walking pace in UK healthcare workers: a nationwide longitudinal cohort study

**DOI:** 10.64898/2026.06.26.26356302

**Authors:** Joel Owen, Christopher A. Martin, Charlotte L Edwardson, Danielle Lamb, Laura J Gray, Michelle Hadjiconstantinou, Emily J Oliver, Charles Goss, Padmasayee Papineni, Holly Reilly, Tom Yates, Katherine Woolf, Manish Pareek

## Abstract

**Objectives:** To examine how self-reported physical activity and walking pace in UK healthcare workers differ ethnicity, migration status, and job role.

**Design:** Prospective cohort study with 6 waves of data collection between 2020 and 2025, analysed using multi-level logistic regression, informed by the UK REACH patient and professional advisory group.

**Setting:** Healthcare settings across the UK. The United Kingdom Research study into Ethnicity And COVID-19 outcomes in Healthcare workers (UK-REACH) cohort.

**Participants:** 18,721 healthcare workers employed in the UK: 28.2% Allied Health Professionals; 75% female; 34% from ethnic minority backgrounds; and 32% born overseas.

**Main outcome measures:** Physical activity and walking pace, as measured by the General Practice Physical Activity Questionnaire (GPPAQ). A ‘Total Physical Activity’ score, and a separate ‘Exercise and Cycling’ score excluding occupational physical activity were derived.

**Results:** Walking pace and physical activity varied significantly by ethnicity, migration status and job role. Compared to White UK-born staff, slower walking pace was reported in ethnic minority staff, with differences greater in staff born overseas (e.g. Asian UK (aOR 0.63, 95%CI 0.56 to 0.71, p<0.001), Asian overseas (aOR 0.38, 95%CI 0.35 to 0.42, p<0.001)). Compared to White UK-born staff, lower odds of reporting being physically active were reported in multiple ethnic groups. Differences were larger for Exercise and Cycling, indicating that occupational physical activity accounted for a large proportion of total physical activity in several groups (e.g. Total Physical Activity: Asian overseas-born (aOR 0.79, 95%CI 0.72 to 0.87, p<0.001); Exercise and Cycling: Asian overseas-born (aOR 0.61, 95%CI 0.56 to 0.67, p=<0.001). Compared to Medical staff, Nursing and Midwifery staff, Pharmacy staff, Dental staff, Other staff, and Healthcare Scientists had significantly lower odds of reporting being physically active. Ambulance staff had higher odds of reporting being physically active when using the Total Physical Activity score (aOR 1.64, 95%CI 1.37 to 1.97 p<0.001), but lower odds for Exercise and Cycling (aOR 0.61 95%CI 0.53 to 0.72, p<0.001).

**Conclusions:** Self-reported physical activity and walking pace vary significantly in UK healthcare workers, by ethnicity, migration status, and job role. Our findings have important implications for understanding workforce health and ethnic health inequalities.

## Background

The World Health Organisation (WHO) defines physical activity as any movement of the body produced by skeletal muscles that requires energy expenditure (1). The benefits of physical activity for health are well-documented (2, 3), with evidence to suggest that increasing both volume and intensity of physical activity bring health benefits (4). For many adults, walking is a common form of physical activity, and walking pace has emerged as a simple and pragmatic indicator of fitness and overall health (5, 6).

Despite the well-established benefits of physical activity, according to self-reported survey data, approximately one-third of the global adult population fail to meet WHO guidance recommending a minimum of 150 minutes of moderate-intensity or 75 minutes of vigorous-intensity PA each week (7). Further, evidence from across multiple settings indicates that physical activity levels vary across a range of demographic factors. In large scale international research for example, females have been found to be less likely to meet physical activity guidelines than males (7). Physical activity levels have also been found to vary by ethnicity and migration status. In the UK for example, White adults have been found to be more active than South Asian adults, but less active than Black adults (8), whilst international research has also identified that migrants report lower levels of physical activity than non-migrants (9). A growing body of research has also explored occupational differences in physical activity levels, with evidence to suggest both that physical activity levels vary across occupations, and that the health effects of occupational versus leisure time physical activity may be different (10, 11). In light of such demographic and occupational differences, calls have been made recently for targeted efforts to increase physical activity in less active groups (7).

As a group whose health has implications for the wellbeing of the wider population (12, 13) Healthcare Workers (HCWs) are an important group to consider physical activity in. The health and wellbeing of the healthcare workforce has implications for patient safety (12), quality of care (13), and retention of staff (14, 15), as well as the sustainability and costs of delivering healthcare services (16). The healthcare workforce is large, with the National Health Service (NHS) in the UK being one of the world’s largest employers (17). Regarding ethnicity, migration status and occupation, the NHS workforce is also highly diverse. Approximately one in four NHS staff members are from ethnically diverse backgrounds, with staff from over 200 countries (18) employed across a wide array of clinical and non-clinical roles (19). Despite this, there is limited evidence to date regarding the amount or intensity of physical activity undertaken by NHS staff, and none that considers how these outcomes vary by ethnicity and migration status. This inhibits the use of evidence-informed targeted policy and intervention to address any inequalities.

To address this gap in the literature we used data from a longitudinal cohort study in HCWs employed in the UK to examine how self-reported physical activity levels and walking pace vary by ethnicity, migration status, and job role, after controlling for age and sex. Our secondary aims were to explore whether any identified variation in our exposures of interest could be accounted for by socio-demographic, health/lifestyle, or shift-related factors.

## Methods

### Overview of the UK-REACH study

For this study we analysed data collected as part of The United Kingdom Research study into Ethnicity And COVID-19 outcomes in Healthcare workers (UK-REACH) cohort study (20). A full description of study methods can be viewed in the cohort study protocol (20) but in brief, UK-REACH is a large national prospective cohort study, established to explore the relationship between ethnicity and COVID-19 related mental and physical health outcomes in HCWs. We gathered electronically at six timepoints between 2020 and 2025. Questionnaire Waves 1-5 followed a single cohort of HCWs, with a further cohort of staff who had joined healthcare since 2021 being added at Wave 6. In the current paper, we present a secondary longitudinal data analysis of UK-REACH physical activity and walking pace data.

#### Participants and recruitment

We invited HCWs (including clinical and non-clinical or ancillary staff) to participate if they were aged 16 or over, working in health and social care in the UK, or registered with one of the UK health regulators. We recruited participants via invitation by UK healthcare regulators, NHS Trusts or healthcare boards. Detailed descriptions of cohort characteristics and recruitments processes are available in cohort profile publications for UK-REACH (21).

#### Outcomes

##### Physical activity level

Physical activity level was measured using the General Practice Physical Activity Questionnaire (GPPAQ) (22). GPPAQ measures physical activity with a composite score combining one item regarding the intensity of physical activity at work, and two items describing the amount of time spent exercising or cycling over the last week (‘Total Physical Activity’). We followed the recommended scoring protocol to use responses to these three items to subsequently classify participants as ‘Inactive’, ‘Moderately Inactive’, ‘Moderately Active’, or ‘Active’. Whilst GPPAQ does also include questions regarding walking, housework/childcare, and gardening/DIY, in keeping with the scoring guidance, we did not include these items in the physical activity score (22). We also created a separate physical activity outcome (‘Exercise and Cycling’) based only on the two items describing the amount of time spent exercising (swimming, jogging, aerobics, gym, football etc) or cycling (including to and from work and during leisure) over the last week (i.e. removing the contribution of PA at work from the Total Physical Activity score). We used the same system to classify participants according to these scores as ‘Inactive’, ‘Moderately Inactive’, ‘Moderately Active’, or ‘Active’. Whilst the GPPAQ scoring protocol uses this 4-point scale, we collapsed both of our physical activity scores into a binary outcome (Active vs Inactive) for analysis.

##### Walking pace

Walking pace is rated via a single question in the GPPAQ (‘How would you describe your usual walking pace?’) using a 4-point scale: ‘Slow pace’, ‘Steady Average Pace’, ‘Brisk Pace’, ‘Fast Pace’. We again collapsed scores into a binary outcome variable for analysis: ‘Slow/Steady’ and ‘Brisk/Fast’ (hereafter ‘Slow’ and ‘Fast’).

#### Exposures

Our primary exposures of interest were self-reported ethnicity and migration status, and job role. These were collected at Wave 1 with participants asked to confirm responses at subsequent waves. Ethnicity was treated as a fixed characteristic, allocated by the first available response and retained throughout. Job role was considered a time-varying variable, where previously reported information was carried forward unless participants stated that it had changed, in which case the new role was inputted. Participants who were newly joined at Wave 6 were asked for the first time then.

For job role we used a list of 54 roles adapted from the NHS Health Careers website (23). These were collapsed for analysis into broader groupings, creating a ten-level categorical variable (For full details see Supplementary Table 1). For ethnicity, we used Office for National Statistics (ONS) 2021 Census categories (24). These were collapsed for analysis into the five broader ethnic groups used by the ONS (White, Asian, Black, Mixed, Other). We created a binary migration status variable by asking participants whether they were born in the UK or overseas. We then formed a 10-level categorical ethnicity by migration status variable: White UK, White Overseas, Asian UK, Asian Overseas, Black UK, Black Overseas, Mixed UK, Mixed Overseas, Other UK, Other Overseas) (For full details see Supplementary Table 1).

#### Covariates

We selected covariates on the basis of existing literature (25, 26) and expert opinion from the research team. Our covariates were sex, age, Index of Multiple Deprivation (IMD) (the official measure of relative deprivation in England (27), expressed in quintiles); smoking and alcohol consumption; number of comorbidities; weekly working hours; and frequency of night shifts. Participants were asked about these variables at wave 1. To minimise demand on participants, subsequent waves asked only whether there were any changes. If no changes were reported, previous responses were carried forward. In the case of changes, the new information replaced what was previously reported from that wave. For details of how these variables were derived, see Supplementary Table 1.

#### Statistical analyses

We used frequency and percentages to describe the cohort in terms of ethnicity, sex, age category and job role. We conducted univariable analyses examining the unadjusted relationship between our outcomes of interest (physical activity and walking pace) and the variables included in our primary (ethnicity, age, sex, and job role) and secondary (IMD quintile, number of comorbidities, alcohol consumption, smoking status, weekly working hours, and night shifts) analyses. Outputs of the univariable analyses (supplementary Tables 6-8) and details regarding how variables were derived (Supplementary Table 1) are presented in the Supplementary Material.

For the multi-variable analyses exploring associations of ethnicity, migration status, or job role, with physical activity or walking pace, we used a complete-case approach, meaning we excluded participants with missing data for any of the covariates included in the relevant analysis. The data followed a two-level structure, with longitudinal observations (Level 1) nested within participants (Level 2). We included participants with one or more data entries for physical activity/walking pace over all survey waves, meaning participants needed only to have completed one wave of data collection to be included. Where participants contributed physical activity or walking pace scores across multiple waves, all were incorporated into the multi-level model. To evaluate the robustness of findings, we performed sensitivity analyses, comparing the results of our main models to those identified when limiting the sample to only those with two or more observations. Details of these analyses are available in Supplementary Figures 1, 2 and 3. All univariable analyses were limited to participants included in the primary analyses.

We used multilevel modelling to maximize data retention, as this approach provides unbiased estimates under the assumption of data being Missing at Random (MAR) and accounts for unbalanced observations per participant. Multilevel modelling was also used to account for the non-independence of observations across their repeated measurements, as we anticipated that a meaningful proportion of the total variance would be attributable to between-participant differences. We tested this assumption by calculating the Intraclass Correlation Coefficient (ICC) which was 0.26 and 0.30 for physical activity and walking pace respectively. To explore the effects of interactions with time, we ran interaction models exploring the interaction between ethnicity and time, and job and time. Wald tests indicated that the interaction terms did not contribute significantly (ethnicity×wave: χ²(35)=20.5, p=0.98; job×wave: χ²(45)=28.07, p=0.98). Interaction terms were therefore not retained in the final models. In our main analyses therefore, we used a random intercept only model to explore the association of (1) ethnicity/migration status, and (2) healthcare profession, with PA levels and walking pace (whilst controlling for age and sex). For each of our main outcomes (‘Total Physical Activity’, ‘Exercise and Cycling’, and ‘Walking Pace’, each expressed as binary variables (Active/Inactive or Fast/Slow)), we first constructed a logistic regression model adjusting for age and sex alongside our main exposures: ethnicity, migration status, and job role (Model 1). We subsequently explored whether any identified relationships could be accounted for by socio-demographic, health/lifestyle, or shift-related variables. As such, we sequentially adjusted our base model for the following sets of variables: Model 2: IMD quintile; Model 3: we further added number of comorbidities, alcohol consumption, smoking status; Model 4: we further added weekly working hours, and frequency of night shifts. We reported our results as adjusted odds ratios (aORs) and 95% confidence intervals (95%CIs) either in Tables or Figures. For all analyses, we used Medical staff and White UK staff as the reference groups.

All analyses were completed via MLWin (using the ‘runmlwin’ command in Stata) and outputs displayed via Stata v.19 (StataCorp. 2025. *Stata Statistical Software: Release 19*. College Station, TX: StataCorp LLC.).

#### Patient and public involvement

The UK-REACH cohort study from which data for the present study was drawn is supported by a patient and professional advisory group. This group meets quarterly, and provides input on all features of the UK-REACH study, such as the content of questionnaires, the burden of taking part, and the dissemination and further research plans. This group also commented on key findings generated for the current paper.

#### Ethics

Ethical approval was obtained for this study via the Health Research Authority (Brighton and Sussex Research Ethics Committee; ethics reference: 20/HRA/4718) and registered with ISRCTN (Reference ISRCTN 11811602). All participants gave informed consent to participate.

## Results

### Cohort description

Demographic details for the cohort are presented in Table 1. The sample was majority female (75.2%), with a mean age of 41 (*SD*=12.3). Approximately one-third (33.9%) were from ethnic minority backgrounds (Asian, Black, Mixed or Other), and approximately one-third (32.3%) were born overseas (White, Asian, Black, Mixed, or Other staff, born overseas). For details of cohort demographics by wave, see Supplementary Table 2. The sample included in these analyses (n=18,271) represented 84.0% of the total UK-REACH cohort (22,290), and was defined by having at least one data point on the primary outcomes (physical activity and walking pace) and exposures (age, sex, ethnicity, and job role). Across the sample the mean number of times an individual completed a questionnaire was two.

**Table 1.** Demographic and occupational details of cohort.

| Variable | n (%)<br>(total n=18,271) |
| --- | --- |
| **Demographic factors** |  |
| **Ethnicity and migration status** |  |
| White UK | 10,322 (56.5) |
| White Overseas | 1,757 (9.6) |
| Asian UK | 1,170 (6.4) |
| Asian Overseas | 2,524 (13.8) |
| Black UK | 253 (1.4) |
| Black Overseas | 995 (5.5) |
| Mixed UK | 519 (2.8) |
| Mixed Overseas | 229 (1.3) |
| Other UK | 117 (0.6) |
| Other Overseas | 385 (2.1) |
| **Sex** |  |
| Male | 4,531 (24.8) |
| Female | 13,740 (75.2) |
| **Age** |  |
| 20 - <30 | 2,078 (11.4) |
| 30 - <40 | 5,374 (29.4) |
| 40 - <50 | 4,255 (23.3) |
| 50 - <60 | 4,023 (22.0) |
| ≥60 | 2,541 (13.9) |
| **Occupational Factors** |  |
| **Healthcare professional group** |  |
| Medical | 3,635 (19.9) |
| Nursing and Midwifery | 4,233 (23.2) |
| Allied Health Professionals (AHPs) (not including scientists) | 5,219 (28.6) |
| Pharmacy | 547 (3.0) |
| Healthcare Scientist | 765 (4.2) |
| Ambulance | 673 (3.7) |
| Dental | 1,203 (6.6) |
| Optical | 375 (2.1) |
| Administrative | 522 (2.9) |
| Other | 1,099 (6.0) |

#### Descriptive Statistics and Univariable Analyses

Unadjusted Odds Ratios for all variables are presented in Supplementary Tables 6, 7, and 8. Full descriptives are presented in Supplementary Tables 3, 4 and 5. Approximately one-third of responding participants at each wave reported being inactive using the total physical activity score (ranging from 36.1% to 42.9%). Using the exercise and cycling only score however, approximately half (47.7% to 57.3%) reported being inactive. Regarding walking pace, just under half of responding participants (ranging from 41.1% to 45.8%) reported being slow/steady walkers across all six waves. Walking pace and activity levels varied across ethnic and occupational groups, as can be seen in Supplementary Tables 3, 4 and 5.

#### Multi-level logistic regression

**FIGURE 1.** aORs and 95% CIs for A. Total GPPAQ score; B. GPPAQ Exercise and Cycling; and C. Walking Pace according to Ethnicity/Migration Status, and Healthcare Profession here.

### Primary analysis

#### Ethnicity and migration status

Forest plots showing aORs and 95% CIs for physical activity and walking pace according to ethnicity and migration status, after controlling for job role, age and sex are shown in Figure 1. Compared to White UK staff, White overseas staff had higher odds of reporting being physically active (aOR 1.18, [95%CI 1.06 – 1.31], p=0.002) when using the total physical activity score. By contrast, lower odds were identified in Asian UK (0.80, [0.70 – 0.90], p<0.001), Asian overseas (0.79, [0.72 – 0.87], p<0.001), Black UK (0.74, [0.58 – 0.95], p=0.018), Black overseas (0.67, [0.58 – 0.77], p<0.001), and Other overseas (0.64, [0.52 – 0.80], p<0.001) staff.

For exercise and cycling only, many of the observed associations became more pronounced with lower odds of reporting being physically active identified in Asian UK (aOR 0.73, [95%CI 0.65 – 0.83], p<0.001), Asian overseas (0.61, [0.56 – 0.67], p=<0.001), and Black overseas (0.46, [0.40 – 0.53], p<0.001) staff. For exercise and cycling, greater evidence was also identified of differences according to migration status (e.g. lower odds of reporting being physically active for Asian Overseas staff than Asian UK staff).

Compared to White UK staff, we identified lower odds of reporting being fast walkers in Asian UK (aOR 0.63, [95%CI 0.56 – 0.71] p<0.001), Asian overseas (0.38, [0.35 – 0.42] p<0.001), Black UK (0.59, [CI 0.46 – 0.76] p=<0.001), Black overseas (0.41, [0.35 – 0.47] p<0.001), Mixed overseas (0.58, [0.44 – 0.75] p<0.001), and Other overseas (0.38, [0.31 – 0.48] p<0.001) staff.

##### Job role

Figure 1 also shows forest plots with aORs and 95% CIs for physical activity and walking pace according to job role, after controlling for ethnicity and migration status, age and sex. Compared to Medical staff, Ambulance staff had higher odds of reporting being physically active using the total physical activity score (aOR 1.64, [95%CI 1.37 – 1.97] p<0.001). By contrast, lower odds were identified in Nursing and Midwifery (0.82, [0.75 – 0.90] p<0.001), Pharmacy (0.74, [0.61 – 0.89] p=0.002), Healthcare Scientists (0.65, [0.56 – 0.75] p=<0.001), Dental (0.80, [0.70 – 0.91] p=0.001), Administrative (0.36, [0.30 – 0.43] p<0.001), and ‘Other’ (0.61, [0.53 – 0.70] p<0.001) staff.

Associations between job role and Exercise and Cycling were generally similar to the associations between job role and total Physical Activity. However, Ambulance staff moved from having the highest odds of reporting being active using the total physical activity score, to having amongst the lowest odds of reporting being physically active when based on exercise and cycling only (aOR 0.61 [95%CI 0.53 – 0.72] p<0.001). Odds of reporting being physically active also fell substantially for Nursing and Midwifery staff (0.49, [0.45 – 0.53] p<0.001) when considering exercise and cycling only.

Compared to Medical staff, lower odds of reporting being fast walkers were identified across all professions, including Nursing and Midwifery (aOR 0.56 [95%CI 0.51 – 0.62] p<0.001), AHPs (0.75, [ 0.69 – 0.82] p<0.001), Pharmacy (0.71, [0.58 – 0.85] p<0.001), Healthcare Scientists (0.68, [0.58 – 0.79] p<0.001), Ambulance (0.46, [0.39 – 0.54] p<0.001), Dental (0.72, [0.63 – 0.82] p<0.001), Optical (0.69, [0.56 – 0.85] p<0.001), Administrative (0.52, [0.43 – 0.62] p<0.001), and Other (0.61, [0.53 – 0.71] p<0.001) staff.

### Secondary analysis

**FIGURE 2.** aORs and 95% CIs for the association between ethnicity/migration status and healthcare profession, with binary Total GPPAQ score. Changes in these relationships following sequential addition of covariates are demonstrated via colour-coded lines.

The relationship between ethnicity and migration status and Total Physical Activity score was largely unchanged following the sequential addition of social deprivation (IMD quintile), health and lifestyle (number of comorbidities, smoking status and alcohol consumption), or work (weekly working hours and frequency of night shifts) covariates. Associations with job role also remained largely unchanged when adjusting for indicators of deprivation (IMD quintile), though they were somewhat attenuated by the subsequent inclusion of a group of health and lifestyle covariates (number of comorbidities, smoking status and alcohol consumption), or work variables (weekly working hours and frequency of night shifts).

**FIGURE 3:** aORs and 95% CIs for the association between ethnicity/migration status and healthcare profession, with binary Exercise and Cycling score. Changes in these relationships following sequential addition of covariates are demonstrated via colour-coded lines.

Adjustment for deprivation indices made little change to the associations between ethnicity and migration status and Exercise and Cycling score. ORs attenuated towards the null after adjustment for health and lifestyle, and shift covariates, but associations with ethnicity and migration status remained significant. Regarding job role, findings were again largely unchanged through the inclusion of social deprivation indicators (IMD quintile). An attenuation towards the null was again identified after adjustment for health and lifestyle, and shift covariates, but the associations with job role remained significant.

**FIGURE 4:** aORs and 95% CIs for the association between ethnicity/migration status and healthcare profession, with binary Walking Pace score. Changes in these relationships following sequential addition of covariates are demonstrated via colour-coded lines.

Some evidence was identified for the relationship between ethnicity and migration status and walking pace being attenuated by the adjustment for health and shift covariates, but overall, the associations were largely unchanged by these sequential additions.

Findings suggested an incremental attenuation of the association between job role and walking pace when adjusting sequentially for deprivation, health, and shift covariates. However, in most cases, the association with job role remained statistically significant.

## Discussion

In this large longitudinal analysis of HCWs working in the UK, we identified that physical activity levels and walking pace varied significantly by ethnicity, migration status, and job role. Most findings remained significant even after controlling for indices of deprivation, health and lifestyle factors, and shift characteristics.

We identified that compared to White UK staff, lower odds of reporting being physically active or having a fast walking pace were reported amongst Asian, Black, and Other staff, with staff in these groups born overseas reporting lower odds than their UK-born counterparts. Regarding physical activity, we found that many of the observed differences were larger when examining Exercise and Cycling activity only. We also observed several significant differences according to job role. Using the Total Physical Activity score, administrative staff had the lowest odds of reporting being physically active, but Nursing and Midwifery staff, Pharmacy staff, Dental staff, Other staff, and Healthcare Scientists also had significantly lower odds of reporting being physically active than Medical staff. Exercise and Cycling scores again provided potentially important insights into how physical activity is accrued differently across the workforce. In the most striking example, Ambulance staff had the highest odds of reporting being active using the Total Physical Activity Score, but amongst the lowest odds when using the Exercise and Cycling score. Similarly, the odds of Nursing and Midwifery staff reporting being active were markedly lower when using the Exercise and Cycling score only, suggesting that these staff groups may be offsetting high occupational activity levels with lower leisure-time physical activity. With evidence to suggest that occupational physical activity is associated with inconsistent or even poor health outcomes (11, 28), these findings may have important implications for understanding workforce health.

### Strengths and Limitations of our study

Our study has several strengths. We have drawn on data from the largest prospective cohort study of HCWs in the UK, with an ethnically and occupationally diverse sample supporting the generalisability of our findings to the wider healthcare workforce. To the best of our knowledge, ours is the only study globally to have reported on how physical activity and walking pace vary by ethnicity and migration status across the healthcare workforce, filling an important gap in the literature given the substantial and growing diversity of the NHS (18). The focus on exploring how these key determinants of health vary by ethnicity aligns closely with core UK government policy aspirations to address drivers of health inequalities (29).

Our study also has several limitations. As with any opt-in cohort study, there is a risk of selection bias. However, across multiple demographic factors (e.g. ethnicity, age, sex) our sample appears to be broadly representative of the wider NHS workforce (30). Relatedly, there is a possibility that healthy participants were more likely to remain in the study. However, our sensitivity analysis comparing findings with a restricted analysis using only participants with two or more observations (Supplementary Figures 1-3) indicated minimal differences. Self-report was used to estimate physical activity and walking pace, and may therefore be susceptible to over-estimation of activity levels (31). However, self-report can be imprecise, particularly in regard to non-structured activities. Questionnaires do not measure ‘small doses’ of physical activity well; these include light-intensity activities, such as folding laundry, and very short bursts of moderate to vigorous-intensity activities, such as running to catch a bus (32). Further research to build on the findings generated here using wearable technologies may be an important next step to explore identified differences in further detail. Finally, the fact that the walking pace item of the GPPAQ does not account for individuals unable to walk represents a further limitation.

### Findings in context

Both physical activity levels and walking pace have well-established connections with multiple aspects of overall health (2, 5, 6). Physical activity has been found to improve a range of health outcomes, including cardiovascular outcomes, reduced mortality rates, and improved mental wellbeing (2, 6, 33), with evidence to suggest that both volume and intensity of physical activity help to drive these outcomes (34). Research has also demonstrated that faster self-reported walking pace is associated with lower mortality rates (35) and is causally linked to reduced cardiovascular disease (36), making it an attractive target for public health messaging (37).

Across our sample, the proportion of staff reporting being physically inactive was broadly comparable with levels reported globally (7). However, this varied by ethnicity, with higher proportions of self-reported physical inactivity in several groups (e.g. Black UK- and Overseas-born staff). Prior work exploring ethnic differences in self-reported physical activity has identified lower levels of physical activity in South Asians than White Europeans (38). Accelerometery data with older adults from the UK Biobank has also found that compared to White participants, lower levels of physical activity were reported in South Asian participants, and higher levels in Black participants (8). In the wider literature regarding walking pace, some evidence has been identified to suggest ethnic differences (39), but evidence is mixed, with other studies finding no such differences (40) or reporting that relationships were accounted for by adjusting for health and lifestyle covariates (41). Notably, existing research in this area for walking pace has relied heavily on older adult samples. In this light, our findings that in this large, working-age sample, ethnic differences in walking pace were evident even after controlling for social deprivation, health and lifestyle factors, and shift covariates are of particular importance.

We adjusted all of our main analyses sequentially for indices of deprivation, health and lifestyle factors, and shift characteristics. We found the associations between our outcomes and ethnicity, migration status, and job role to be slightly attenuated by these adjustments in some cases, meaning efforts to address co-morbidity management or behaviours related to smoking and alcohol consumption may reduce some of the ethnic and migrant inequalities observed. However, our findings demonstrate that in this large sample of UK HCWs, ethnicity, migration status and job role are meaningful predictors of both physical activity levels and walking pace, even after controlling for other important covariates. More research is therefore urgently needed to uncover the drivers of these differences.

### Implications and future research

The health and wellbeing of the healthcare workforce has significant implications for the effective delivery of NHS services. Poor staff wellbeing is costly (16) and effects both patient care and staff retention (12–14). Workplace absence in the NHS is high, with sickness rates over double that of the wider UK workforce (42). With physical activity and walking pace both known to improve a variety of health outcomes (2, 3, 5) and walking pace a powerful prognostic indicator of cardiovascular mortality (43), the differences we have identified in these outcomes according to ethnicity, migration status, and job role in this large cohort of NHS staff are important. Our findings may point towards workforce groups particularly at risk of poor health outcomes. Recent calls have been made to use walking pace to identify individuals at risk of ill-health (6), and our findings suggest that efforts to incorporate this into workforce health monitoring may be warranted. Secondly, recent evidence suggests that leisure-time and transport-related physical activity are positively associated with mental health, whilst occupational physical activity is positively associated with mental *ill*-health (11, 44). As such, our finding that staff groups such as Ambulance and Nursing staff may be offsetting high-levels of occupational physical activity with lower physical activity levels outside of work, suggest that occupational factors may contribute to health inequalities in the workforce. Finally, with even modest increases in physical activity reported to lead to meaningful health benefits (3), our findings suggest that physical activity interventions for healthcare staff may be important for employers and policy-makers interested in workforce wellbeing. Targeted interventions to increase physical activity and walking pace amongst staff with particularly low physical activity levels, slow walking pace, or low proportions of physical activity undertaken outside work may be of particular value, and may help contribute towards efforts to reduce health inequalities in the workforce.

## Conclusions

Using data from the largest known cohort study of HCWs to date, we have identified significant variation in physical activity levels and walking pace in HCWs according to ethnicity, migration status, and job role. In most cases, associations remained significant even after controlling for social deprivation, health and lifestyle factors, and shift covariates, highlighting the importance of targeted and tailored workforce-focused interventions alongside work on broader social determinants of health. These findings have important implications for understanding, as well as generating evidence-informed intervention to address, workforce activity, health and health inequalities.

## What is already known on this topic

- Physical activity is known to improve both mental and physical health, though evidence suggests that leisure and travel time physical activity may be better for health than occupational physical activity
- Walking pace is considered a key indicator of overall fitness and health, with faster walking pace associated with multiple health benefits including reduced mortality and cardiovascular disease
- Healthcare workers experience high levels of mental and physical ill-health, and typically report either low levels of physical activity, or report accruing much of their physical activity through work

## What this study adds

- Significant variation in physical activity levels and walking pace amongst UK healthcare workers was identified according to ethnicity, migration status and job role
- Several ethnic (e.g. Asian overseas-born staff) and occupational (e.g. Ambulance staff) staff groups report undertaking a relatively higher proportion of their total physical activity at work, with important implications for workforce health

## Data Availability

The data used for this study are available upon request.

## Acknowledgements

This study was supported by the National Institute for Health and Care Research (NIHR) Leicester BRC. The views expressed are those of the author(s) and not necessarily those of the NIHR or the Department of Health and Social Care. LG is funded by the National Institute for Health and Care Research (NIHR) Applied Research Collaboration East Midlands (ARC EM) and Leicester NIHR Biomedical Research Centre (BRC (NIHR203327)). LG is an NIHR Senior Investigator and a member of the NIHR Multiple Long-Term Conditions Cross-NIHR Collaboration. The views expressed are those of the author(s) and not necessarily those of the NIHR or the Department of Health and Social Care. LG is supported by British Heart Foundation Research Excellence Award (RE/24/130031).

DL was funded by the National Institute for Health and Care Research Applied Research Collaboration North Thames. The views expressed are those of the author(s) and not necessarily those of the NIHR or the Department of Health and Social Care.

## Data sharing statement

The data used for this study are available upon request.

## Transparency statement

The corresponding author affirms that this manuscript is an honest, accurate, and transparent account of the study being reported; that no important aspects of the study have been omitted; and that any discrepancies from the study as planned have been explained.

## Contributor and guarantor information

MP is the guarantor of this work. MP led the conceptualisation of the overarching UK-REACH study from which the present data was drawn. The conceptualisation of the present study was led by MP, KW, CM and JO. Analysis was conducted by CM and JO. Writing of the original draft was led by JO and CM. All authors contributed to interpretation and presentation of results through reviewing and commenting on drafts and the final submission. The corresponding author attests that all listed authors meet authorship criteria and that no others meeting the criteria have been omitted.

