## Supplementary Material for "Demographic and occupational differences in physical activity and walking pace in UK healthcare workers: a nationwide longitudinal cohort study"

**Content**

Supplementary Tables 1 – 8

Supplementary Figures 1-3

**Supplementary Table 1. Derivation of covariates from questionnaire data**

| Variable | Description |
|---|---|
| **Age** | Continuous variable. Age in years. Derived from date of birth entered by participants at registration. |
| **Sex** | Binary variable. Participants were asked their sex assigned at birth. |
| **Ethnicity** | Categorical variable. Participants were asked to select their ethnicity from a list of the 18 Office for National Statistics categories:<br>Asian/Asian British – Indian<br>Asian/Asian British – Pakistani<br>Asian/Asian British – Bangladeshi<br>Asian/Asian British – Chinese<br>Asian/Asian British - Any other Asian background<br>Black/African/Caribbean/Black British - African<br>Black/African/Caribbean/Black British – Caribbean<br>Black/African/Caribbean/Black British - Any other Black/African/Caribbean background<br>Mixed/Multiple ethnic groups - White and Black Caribbean<br>Mixed/Multiple ethnic groups - White and Black African<br>Mixed/Multiple ethnic groups - White and Asian<br>Mixed/Multiple ethnic groups - Any other Mixed/multiple ethnic background<br>White - English/Welsh/Scottish/Northern Irish/British<br>White – Irish<br>White - Gypsy or Irish Traveller<br>White - Any other white background<br>Other ethnic group – Arab<br>Other ethnic group - Any other ethnic background<br><br>These were categorised into the 5 broader Office for National Statistics ethnicity categories (Asian, Black, Mixed, White, Other). |

| | |
|---|---|
| **Migration status** | Binary variable. Participants were asked whether they were born in the UK. |
| **Occupation** | Categorical variable. Participants were asked to select their main job/role. Categorised as below:<br><br>**Doctor or medical support** - Doctor, Advanced Critical Care Practitioner, Anaesthesia associate, Surgical Care Practitioner, Other medical associate<br>**Nurse, NA or Midwife -** Advanced Nurse Practitioner, Healthcare assistant, Maternity support worker, Midwife, Nurse, Nursing Associate, Other nursing and midwifery role,<br>**Allied Health Professional (including pharmacists, ambulance workers and those in optical roles)** - Arts therapist, Biomedical scientist, Chiropodist/Podiatrist, Clinical scientist, Dietician, Hearing aid dispenser, Occupational therapist, Operating department practitioner, Orthoptist, Physiotherapist, Practitioner psychologist, Prosthetist / Orthotist, Radiographer, Speech and language therapist, Other Allied Health Professional role, Emergency medical, Paramedic , Other ambulance role, OT Support , Phlebotomist, Physiotherapy Assistant, Radiography Other clinical support role , Pharmacist , Pharmacy technician, Other pharmacy role, Optical - Dispensing optician, Optometrist, Other Optical role<br>**Dental -** Clinical dental technician, Dental Hygienist, Dental nurse, Dental technician, Dentist, Other dental role<br>**Admin, estates or other –** Administration, Catering services, Domestic services, Estates services, Porter, Other |
| **Index of Multiple Deprivation (IMD) quintile** | Ordinal variable. Participants provided their residential postcode on registration for the study. This was used to determine the Index of Multiple Deprivation (the official measure of deprivation for small areas of England) in the area in which they live. The IMD ranks all areas in England based on 7 measures of deprivation and the ranks can be expressed as quintiles. Lower quintiles indicate more deprivation. |
| **Comorbidities** | Categorical variable. Participants were asked to select comorbidities from the following list:<br><br>Organ transplant \| Diabetes (Type I or II) \| Heart disease or heart problems \| Hypertension \| Overweight \| Stroke \| Kidney disease \| Liver disease \| Anaemia \| Asthma \| Other lung condition such as COPD, bronchitis or emphysema \| Cancer \| Condition affecting the brain and nerves (e.g. Dementia, Parkinson's, Multiple Sclerosis) \| A weakened immune system or reduced ability to deal with infections (as a result of a disease or treatment) \| Depression \| Anxiety \| Psychiatric disorder \| Prefer not to answer |

| | |
|---|---|
| | Number of comorbidities was generated by taking first reported number of comorbidities for each participant, and adding to this any new comorbidities reported later. |
| **Smoking status** | Binary variable. Participants were asked to indicate their current smoking status. Never and ex-smokers were grouped together and compared with current smokers. |
| **Alcohol use** | Categorical variable. Participants were asked to indicate how often they drank alcohol using the following categories: "Never", "Monthly or less", "2-4 times per month", "2-3 times per week", "4+ times per week", or "Prefer not to answer" |
| **Night shift pattern** | Categorical variable. Participants were asked how often they work night shifts, using the following categories: “Never”, “Less than once a month”, “Once a month or more, but not every week”, “Once a week or more, but not every shift”, or “I always work nights”. |
| **Work hours** | Categorical variable. Participants were asked to report how many hours they work in a week, and responses were then categorised as either "0–20 hours", "21–30 hours", "31–40 hours", or "More than 40 hours" |

**Supplementary Table 2. Demographic information by wave**

| Variable | Wave 1<br>N (%) | Wave 2<br>N (%) | Wave 3<br>N (%) | Wave 4<br>N (%) | Wave 5<br>N (%) | Wave 6<br>N (%) |
|---|---|---|---|---|---|---|
| **White UK** | 7,467 (60.63) | 3,386 (64.90) | 2,832 (65.48) | 1,950 (68.83) | 2,740 (68.19) | 4,870 (53.39) |
| **White overseas** | 1,060 (8.61) | 449 (8.61) | 379 (8.76) | 227 (8.01) | 306 (7.62) | 941 (10.32) |
| **Asian UK** | 856 (6.95) | 377 (7.23) | 303 (7.01) | 193 (6.81) | 226 (5.62) | 523 (5.73) |
| **Asian overseas** | 1,450 (11.77) | 512 (9.81) | 397 (9.18) | 227 (8.01) | 337 (8.39) | 1,333 (14.61) |
| **Black UK** | 150 (1.22) | 54 (1.04) | 53 (1.23) | 34 (1.20) | 57 (1.42) | 149 (1.63) |
| **Black Overseas** | 365 (2.96) | 114 (2.19) | 92 (2.13) | 47 (1.66) | 76 (1.89) | 685 (7.51) |
| **Mixed UK** | 377 (3.06) | 154 (2.95) | 137 (3.17) | 92 (3.25) | 127 (3.16) | 244 (2.67) |
| **Mixed Overseas** | 132 (1.07) | 52 (1.00) | 42 (0.97) | 18 (0.64) | 41 (1.02) | 115 (1.26) |
| **Other UK** | 52 (0.42) | 26 (0.50) | 17 (0.39) | 8 (0.28) | 13 (0.32) | 48 (0.53) |
| **Other Overseas** | 184 (1.49) | 78 (1.50) | 55 (1.27) | 32 (1.13) | 43 (1.07) | 172 (1.89) |

| | | | | | | |
|---|---|---|---|---|---|---|
| **Total** | 12,316 (100.00) | 5,217 (100.00) | 4,325 (100.00) | 2,833 (100.00) | 4,018 (100.00) | 9,122 (100.00) |
| **Medical Staff** | 2,845 (22.69) | 1,298 (24.26) | 1,051 (23.81) | 686 (24.10) | 910 (22.74) | 1,508 (16.09) |
| **Nurses, NAs, Midwives** | 2,556 (20.38) | 1,103 (20.62) | 948 (21.48) | 611 (21.47) | 922 (23.04) | 2,410 (25.72) |
| **AHPs (not including scientists)** | 3,732 (29.76) | 1,514 (28.30) | 1,254 (28.41) | 837 (29.41) | 1,184 (29.59) | 2,467 (26.33) |
| **Pharmacy** | 244 (1.95) | 99 (1.85) | 89 (2.02) | 53 (1.86) | 69 (1.72) | 357 (3.81) |
| **Healthcare Scientist** | 553 (4.41) | 248 (4.64) | 223 (5.05) | 133 (4.67) | 164 (4.10) | 372 (3.97) |
| **Ambulance** | 443 (3.53) | 174 (3.25) | 159 (3.60) | 93 (3.27) | 147 (3.67) | 337 (3.60) |
| **Dental** | 758 (6.04) | 304 (5.68) | 234 (5.30) | 154 (5.41) | 213 (5.32) | 604 (6.45) |
| **Optical** | 297 (2.37) | 141 (2.64) | 96 (2.17) | 53 (1.86) | 87 (2.17) | 133 (1.42) |
| **Administrative** | 241 (1.92) | 132 (2.47) | 95 (2.15) | 74 (2.60) | 90 (2.25) | 361 (3.85) |
| **Other** | 424 (3.38) | 189 (3.53) | 158 (3.58) | 134 (4.71) | 180 (4.50) | 531 (5.67) |
| **Total** | 12,541 (100.00) | 5,350 (100.00) | 4,414 (100.00) | 2,846 (100.00) | 4,002 (100.00) | 9,370 (100.00) |
| **Male** | 2,857 (23.58) | 1,287 (24.72) | 1,061 (24.62) | 710 (25.09) | 979 (24.67) | 2,397 (26.31) |
| **Female** | 9,236 (76.23) | 3,915 (75.20) | 3,246 (75.31) | 2,118 (74.84) | 2,987 (75.26) | 6,683 (73.34) |
| **Missing** | 23 (0.19) | 4 (0.08) | 3 (0.07) | 2 (0.07) | 3 (0.08) | 32 (0.35) |
| **Total** | 12,116 (100) | 5,206 (100) | 4,310 (100) | 2,830 (100) | 3,969 (100) | 9,112 (100) |
| **20 - <30 years** | 780 (6.42) | 268 (5.13) | 206 (4.75) | 107 (3.71) | 138 (3.46) | 1,395 (14.83) |
| **30 - <40 years** | 2,837 (23.34) | 1,189 (22.76) | 964 (22.23) | 565 (19.58) | 751 (18.86) | 3,076 (32.70) |
| **40 - <50 years** | 2,990 (24.60) | 1,237 (23.68) | 1,025 (23.64) | 661 (22.90) | 886 (22.24) | 1,902 (20.22) |
| **50 - <60 years** | 3,293 (27.10) | 1,449 (27.74) | 1,247 (28.76) | 858 (29.73) | 1,257 (31.56) | 1,694 (18.01) |
| **>60 years** | 2,192 (18.03) | 1,059 (20.27) | 865 (19.95) | 637 (22.07) | 934 (23.45) | 1,013 (10.77) |
| **Missing** | 60 (0.49) | 22 (0.42) | 29 (0.67) | 58 (2.01) | 17 (0.43) | 328 (3.49) |
| **Total** | 12,153 (100) | 5,224 (100) | 4,336 (100) | 2,886 (100) | 3,983 (100) | 9,408 (100) |
| **IMD 1** | 1,001 (8.28) | 373 (7.17) | 293 (6.80) | 183 (6.47) | 270 (6.81) | 1,123 (12.37) |
| **IMD 2** | 1,759 (14.55) | 769 (14.78) | 639 (14.84) | 408 (14.43) | 548 (13.82) | 1,596 (17.58) |
| **IMD 3** | 2,201 (18.20) | 956 (18.38) | 794 (18.44) | 509 (18.00) | 694 (17.50) | 1,710 (18.83) |

| | | | | | | |
|---|---|---|---|---|---|---|
| **IMD 4** | 2,615 (21.62) | 1,162 (22.34) | 963 (22.36) | 635 (22.45) | 901 (22.72) | 1,804 (19.87) |
| **IMD 5** | 3,123 (25.82) | 1,364 (26.22) | 1,131 (26.26) | 739 (26.13) | 1,079 (27.21) | 1,877 (20.67) |
| **Missing** | 1,394 (11.53) | 578 (11.11) | 487 (11.31) | 354 (12.52) | 474 (11.95) | 970 (10.68) |
| **Total** | 12,093 | 5,202 | 4,307 | 2,828 | 3,966 | 9,080 |
| **Alcohol Never** | 2,042 (16.89) | 854 (16.42) | 708 (16.44) | 467 (16.51) | 621 (15.66) | 2,476 (27.27) |
| **Monthly or less** | 2,999 (24.80) | 1,237 (23.78) | 1,035 (24.03) | 710 (25.11) | 977 (24.63) | 2,765 (30.45) |
| **2-4 x month** | 2,871 (23.74) | 1,184 (22.76) | 992 (23.03) | 636 (22.49) | 899 (22.67) | 2,002 (22.05) |
| **2-3 x week** | 2,897 (23.96) | 1,273 (24.47) | 1,033 (23.98) | 670 (23.69) | 942 (23.75) | 1,342 (14.78) |
| **4+ x week** | 1,244 (10.29) | 520 (10.00) | 444 (10.31) | 298 (10.54) | 415 (10.46) | 475 (5.23) |
| **Missing** | 40 (0.33) | 134 (2.58) | 95 (2.21) | 47 (1.66) | 112 (2.82) | 20 (0.22) |
| **Total** | 12,093 (100) | 5,202 (100) | 4,307 (100) | 2,828 (100) | 3,966 (100) | 9,080 (100) |
| **Smoking ever** | 3,208 (26.53) | 1,289 (24.78) | 1,048 (24.33) | 658 (23.27) | 934 (23.55) | 2,265 (24.94) |
| **Smoking never** | 8,783 (72.63) | 3,756 (72.20) | 3,147 (73.07) | 2,116 (74.82) | 2,906 (73.27) | 6,723 (74.04) |
| **Missing** | 102 (0.84) | 157 (3.02) | 112 (2.60) | 54 (1.91) | 126 (3.18) | 92 (1.01) |
| **Total** | 12,093 (100) | 5,202 (100) | 4,307 (100) | 2,828 (100) | 3,966 (100) | 9,080 (100) |
| **Working hrs 0-20** | 1,162 (9.61) | 514 (9.88) | 426 (9.89) | 312 (11.03) | 418 (10.54) | 806 (8.88) |
| **21-30 hrs** | 2,281 (18.86) | 1,013 (19.47) | 818 (18.99) | 561 (19.84) | 774 (19.52) | 1,474 (16.23) |
| **31-40 hrs** | 6,021 (49.79) | 2,373 (45.62) | 1,918 (44.53) | 1,178 (41.65) | 1,578 (39.79) | 5,386 (59.32) |
| **40+ hrs** | 1,931 (15.97) | 862 (16.57) | 668 (15.51) | 415 (14.67) | 519 (13.09) | 1,202 (13.24) |
| **Missing** | 698 (5.77) | 440 (8.46) | 477 (11.07) | 362 (12.80) | 677 (17.07) | 212 (2.33) |
| **Total** | 12,093 (100) | 5,202 (100) | 4,307 (100) | 2,828 (100) | 3,966 (100) | 9,080 (100) |
| **Nights Never** | 8,639 (71.44) | 2,362 (45.41) | 1,920 (44.58) | 1,261 (44.59) | 1,671 (42.13) | 6,350 (69.93) |
| **<1 x month** | 601 (4.97) | 275 (5.29) | 231 (5.36) | 145 (5.13) | 191 (4.82) | 508 (5.59) |
| **≥ 1 x month, but not every week** | 1,155 (9.55) | 491 (9.44) | 374 (8.68) | 240 (8.49) | 292 (7.36) | 1,049 (11.55) |

| | | | | | | |
|---|---|---|---|---|---|---|
| **≥ 1 x week, but not every shift** | 832 (6.88) | 302 (5.81) | 233 (5.41) | 150 (5.30) | 190 (4.79) | 839 (9.24) |
| **Nights always** | 185 (1.53) | 44 (0.85) | 36 (0.84) | 24 (0.85) | 38 (0.96) | 189 (2.08) |
| **Missing** | 681 (5.63) | 522 (10.03) | 517 (12.00) | 349 (12.34) | 689 (17.37) | 145 (1.60) |
| **Total** | 12,093 (100) | 5,202 (100) | 4,307 (100) | 2,828 (100) | 3,966 (100) | 9,080 (100) |

**Supplementary Table 3. Frequency and percentages of Inactive vs Active staff by wave, when using total GPPAQ score**

| | Wave 1 | | Wave 2 | | Wave 3 | | Wave 4 | | Wave 5 | | Wave 6 | |
|---|---|---|---|---|---|---|---|---|---|---|---|---|
| | Inactive N (%) | Active N (%) | Inactive N (%) | Active N (%) | Inactive N (%) | Active N (%) | Inactive N (%) | Active N (%) | Inactive N (%) | Active N (%) | Inactive N (%) | Active N (%) |
| **White UK** | 2,964 (42.99) | 3,931 (57.01) | 1,236 (39.41) | 1,900 (60.59) | 1,021 (39.38) | 1,572 (60.62) | 695 (39.56) | 1,062 (60.44) | 928 (37.43) | 1,551 (62.57) | 1,527 (34.87) | 2,852 (65.13) |
| **White overseas** | 372 (37.42) | 622 (62.58) | 149 (35.99) | 265 (64.01) | 123 (34.94) | 229 (65.06) | 70 (34.65) | 132 (65.35) | 89 (31.67) | 192 (68.33) | 258 (29.38) | 620 (70.62) |
| **Asian UK** | 357 (44.46) | 446 (55.54) | 152 (43.18) | 200 (56.82) | 106 (37.32) | 178 (62.68) | 75 (41.21) | 107 (58.79) | 82 (37.96) | 134 (62.04) | 183 (36.38) | 320 (63.62) |
| **Asian overseas** | 570 (42.82) | 761 (57.18) | 168 (35.82) | 301 (64.18) | 144 (38.71) | 228 (61.29) | 85 (40.09) | 127 (59.91) | 125 (40.72) | 182 (59.28) | 488 (39.80) | 738 (60.20) |
| **Black UK** | 67 (48.55) | 71 (51.45) | 25 (48.08) | 27 (51.92) | 21 (41.18) | 30 (58.82) | 16 (53.33) | 14 (46.67) | 20 (38.46) | 32 (61.54) | 58 (42.34) | 79 (57.66) |
| **Black Overseas** | 175 (50.87) | 169 (49.13) | 47 (45.63) | 56 (54.37) | 36 (46.15) | 42 (53.85) | 17 (40.48) | 25 (59.52) | 27 (38.57) | 43 (61.43) | 277 (43.62) | 358 (56.38) |
| **Mixed UK** | 132 (37.61) | 219 (62.39) | 57 (39.31) | 88 (60.69) | 53 (41.73) | 74 (58.27) | 29 (34.12) | 56 (65.88) | 40 (32.79) | 82 (67.21) | 88 (37.61) | 146 (62.39) |
| **Mixed Overseas** | 56 (45.16) | 68 (54.84) | 23 (46.00) | 27 (54.00) | 17 (41.46) | 24 (58.54) | 8 (61.54) | 5 (38.46) | 16 (42.11) | 22 (57.89) | 35 (33.33) | 70 (66.67) |
| **Other UK** | 25 (49.02) | 26 (50.98) | 5 (19.23) | 21 (80.77) | 4 (23.53) | 13 (76.47) | 1 (12.50) | 7 (87.50) | 4 (33.33) | 8 (66.67) | 18 (38.30) | 29 (61.70) |
| **Other Overseas** | 90 (50.56) | 88 (49.44) | 36 (48.65) | 38 (51.35) | 27 (50.94) | 26 (49.06) | 16 (50.00) | 16 (50.00) | 20 (48.78) | 21 (51.22) | 63 (38.89) | 99 (61.11) |
| **Total** | 4,808 (42.89) | 6,401 (57.11) | 1,898 (39.37) | 2,923 (60.63) | 1,552 (39.11) | 2,416 (60.89) | 1,012 (39.48) | 1,551 (60.52) | 1,351 (37.34) | 2,267 (62.66) | 2,995 (36.06) | 5,311 (63.94) |
| **Medical Staff** | 1,041 (39.27) | 1,610 (60.73) | 423 (35.16) | 780 (64.84) | 353 (36.32) | 619 (63.68) | 227 (36.09) | 402 (63.91) | 287 (34.70) | 540 (65.30) | 484 (36.17) | 854 (63.83) |
| **Nurses, NAs, Midwives** | 1,148 (48.26) | 1,231 (51.74) | 459 (44.52) | 572 (55.48) | 379 (42.49) | 513 (57.51) | 244 (43.65) | 315 (56.35) | 355 (43.35) | 464 (56.65) | 830 (37.57) | 1,379 (62.43) |

| | | | | | | | | | | | | |
|---|---|---|---|---|---|---|---|---|---|---|---|---|
| **AHPs (not including scientists)** | 1,311 (38.26) | 2,116 (61.74) | 508 (36.73) | 875 (63.27) | 397 (35.13) | 733 (64.87) | 275 (36.72) | 474 (63.28) | 348 (31.96) | 741 (68.04) | 703 (31.27) | 1,545 (68.73) |
| **Pharmacy** | 122 (52.59) | 110 (47.41) | 41 (44.57) | 51 (55.43) | 33 (38.82) | 52 (61.18) | 20 (40.00) | 30 (60.00) | 27 (40.91) | 39 (59.09) | 127 (37.35) | 213 (62.65) |
| **Healthcare Scientist** | 281 (53.52) | 244 (46.48) | 104 (43.88) | 133 (56.12) | 97 (45.33) | 117 (54.67) | 49 (41.18) | 70 (58.82) | 70 (47.62) | 77 (52.38) | 141 (41.11) | 202 (58.89) |
| **Ambulance** | 101 (23.88) | 322 (76.12) | 39 (23.64) | 126 (76.36) | 40 (27.21) | 107 (72.79) | 20 (24.10) | 63 (75.90) | 34 (23.45) | 111 (76.55) | 57 (18.27) | 255 (81.73) |
| **Dental** | 321 (46.12) | 375 (53.88) | 114 (39.86) | 172 (60.14) | 96 (44.65) | 119 (55.35) | 60 (42.25) | 82 (57.75) | 74 (38.54) | 118 (61.46) | 190 (34.67) | 358 (65.33) |
| **Optical** | 125 (46.82) | 142 (53.18) | 50 (39.68) | 76 (60.32) | 31 (36.47) | 54 (63.53) | 17 (36.96) | 29 (63.04) | 32 (39.51) | 49 (60.49) | 43 (37.07) | 73 (62.93) |
| **Administrative** | 143 (64.41) | 79 (35.59) | 71 (56.35) | 55 (43.65) | 55 (62.50) | 33 (37.50) | 40 (57.14) | 30 (42.86) | 46 (52.27) | 42 (47.73) | 210 (60.69) | 136 (39.31) |
| **Other** | 215 (55.56) | 172 (44.44) | 89 (51.74) | 83 (48.26) | 71 (50.71) | 69 (49.29) | 60 (51.72) | 56 (48.28) | 78 (47.56) | 86 (52.44) | 210 (41.50) | 296 (58.50) |
| **Total** | 4,808 (42.89) | 6,401 (57.11) | 1,898 (39.37) | 2,923 (60.63) | 1,552 (39.11) | 2,416 (60.89) | 1,012 (39.48) | 1,551 (60.52) | 1,351 (37.34) | 2,267 (62.66) | 2,995 (36.06) | 5,311 (63.94) |

**Supplementary Table 4. Frequency and percentages of Inactive vs Active staff by wave, when using Exercise and Cycling only score**

| | **Wave 1** | | **Wave 2** | | **Wave 3** | | **Wave 4** | | **Wave 5** | | **Wave 6** | |
|---|---|---|---|---|---|---|---|---|---|---|---|---|
| | Inactive N (%) | Active N (%) | Inactive N (%) | Active N (%) | Inactive N (%) | Active N (%) | Inactive N (%) | Active N (%) | Inactive N (%) | Active N (%) | Inactive N (%) | Active N (%) |

| | | | | | | | | | | | | |
|---|---|---|---|---|---|---|---|---|---|---|---|---|
| **White UK** | 3,998 (56.61) | 3,064 (43.39) | 1,627 (50.29) | 1,608 (49.71) | 553 (50.73) | 537 (49.27) | 383 (51.62) | 359 (48.38) | 1,286 (48.24) | 1,380 (51.76) | 2,324 (48.82) | 2,436 (51.18) |
| **White overseas** | 546 (53.58) | 473 (46.42) | 192 (44.24) | 242 (55.76) | 71 (43.03) | 94 (56.97) | 39 (44.83) | 48 (55.17) | 122 (40.40) | 180 (59.60) | 386 (41.42) | 546 (58.58) |
| **Asian UK** | 475 (57.93) | 345 (42.07) | 202 (54.59) | 168 (45.41) | 67 (48.55) | 71 (51.45) | 39 (48.75) | 41 (51.25) | 102 (45.74) | 121 (54.26) | 250 (48.64) | 264 (51.36) |
| **Asian overseas** | 812 (59.05) | 563 (40.95) | 243 (49.19) | 251 (50.81) | 91 (56.88) | 69 (43.12) | 43 (50.00) | 43 (50.00) | 161 (50.16) | 160 (49.84) | 778 (59.16) | 537 (40.84) |
| **Black UK** | 84 (58.74) | 59 (41.26) | 27 (50.94) | 26 (49.06) | 12 (40.00) | 18 (60.00) | 13 (61.90) | 8 (38.10) | 25 (45.45) | 30 (54.55) | 75 (51.72) | 70 (48.28) |
| **Black Overseas** | 249 (70.34) | 105 (29.66) | 65 (58.04) | 47 (41.96) | 26 (57.78) | 19 (42.22) | 10 (47.62) | 11 (52.38) | 44 (59.46) | 30 (40.54) | 456 (68.26) | 212 (31.74) |
| **Mixed UK** | 193 (53.91) | 165 (46.09) | 70 (47.95) | 76 (52.05) | 36 (52.94) | 32 (47.06) | 16 (38.10) | 26 (61.90) | 52 (41.94) | 72 (58.06) | 117 (48.35) | 125 (51.65) |
| **Mixed Overseas** | 74 (58.27) | 53 (41.73) | 28 (52.83) | 25 (47.17) | 12 (52.17) | 11 (47.83) | 3 (50.00) | 3 (50.00) | 20 (50.00) | 20 (50.00) | 59 (52.68) | 53 (47.32) |
| **Other UK** | 30 (58.82) | 21 (41.18) | 6 (23.08) | 20 (76.92) | 2 (40.00) | 3 (60.00) | 1 (33.33) | 2 (66.67) | 4 (30.77) | 9 (69.23) | 23 (47.92) | 25 (52.08) |
| **Other Overseas** | 121 (66.48) | 61 (33.52) | 48 (62.34) | 29 (37.66) | 17 (73.91) | 6 (26.09) | 5 (45.45) | 6 (54.55) | 27 (64.29) | 15 (35.71) | 87 (51.48) | 82 (48.52) |
| **Total** | 6,582 (57.28) | 4,909 (42.72) | 2,508 (50.16) | 2,492 (49.84) | 887 (50.77) | 860 (49.23) | 552 (50.23) | 547 (49.77) | 1,843 (47.75) | 2,017 (52.25) | 4,555 (51.15) | 4,350 (48.85) |
| **Medical Staff** | 1,359 (49.93) | 1,363 (50.07) | 549 (43.78) | 705 (56.22) | 195 (49.12) | 202 (50.88) | 102 (40.80) | 148 (59.20) | 366 (41.50) | 516 (58.50) | 659 (44.50) | 822 (55.50) |
| **Nurses, NAs, Midwives** | 1,665 (68.35) | 771 (31.65) | 617 (58.04) | 446 (41.96) | 222 (54.81) | 183 (45.19) | 149 (59.60) | 101 (40.40) | 507 (56.52) | 390 (43.48) | 1,460 (61.63) | 909 (38.37) |
| **AHPs (not including scientists)** | 1,877 (53.43) | 1,636 (46.57) | 697 (48.37) | 744 (51.63) | 238 (46.12) | 278 (53.88) | 157 (48.01) | 170 (51.99) | 503 (43.63) | 650 (56.37) | 1,102 (45.69) | 1,310 (54.31) |
| **Pharmacy** | 150 (63.29) | 87 (36.71) | 56 (58.33) | 40 (41.67) | 24 (66.67) | 12 (33.33) | 8 (50.00) | 8 (50.00) | 32 (46.38) | 37 (53.62) | 170 (48.30) | 182 (51.70) |

| | | | | | | | | | | | | |
|---|---|---|---|---|---|---|---|---|---|---|---|---|
| **Healthcare Scientist** | 336 (63.04) | 197 (36.96) | 131 (53.47) | 114 (46.53) | 49 (55.06) | 40 (44.94) | 22 (55.00) | 18 (45.00) | 86 (54.09) | 73 (45.91) | 187 (50.95) | 180 (49.05) |
| **Ambulance** | 234 (54.55) | 195 (45.45) | 84 (50.00) | 84 (50.00) | 32 (43.24) | 42 (56.76) | 29 (61.70) | 18 (38.30) | 79 (54.11) | 67 (45.89) | 156 (46.71) | 178 (53.29) |
| **Dental** | 412 (57.54) | 304 (42.46) | 138 (47.10) | 155 (52.90) | 48 (53.33) | 42 (46.67) | 23 (47.92) | 25 (52.08) | 102 (48.80) | 107 (51.20) | 274 (46.92) | 310 (53.08) |
| **Optical** | 153 (54.45) | 128 (45.55) | 66 (48.18) | 71 (51.82) | 15 (48.39) | 16 (51.61) | 6 (37.50) | 10 (62.50) | 36 (41.86) | 50 (58.14) | 57 (43.18) | 75 (56.82) |
| **Administrative** | 149 (66.82) | 74 (33.18) | 72 (56.69) | 55 (43.31) | 29 (65.91) | 15 (34.09) | 22 (57.89) | 16 (42.11) | 48 (54.55) | 40 (45.45) | 223 (63.35) | 129 (36.65) |
| **Other** | 247 (61.60) | 154 (38.40) | 98 (55.68) | 78 (44.32) | 35 (53.85) | 30 (46.15) | 34 (50.75) | 33 (49.25) | 84 (49.12) | 87 (50.88) | 267 (51.15) | 255 (48.85) |
| **Total** | 6,582 (57.28) | 4,909 (42.72) | 2,508 (50.16) | 2,492 (49.84) | 887 (50.77) | 860 (49.23) | 552 (50.23) | 547 (49.77) | 1,843 (47.75) | 2,017 (52.25) | 4,555 (51.15) | 4,350 (48.85) |

**Supplementary Table 5. Frequency and percentages of Slow vs Fast Walking Pace by wave**

| | **Wave 1** | | **Wave 2** | | **Wave 3** | | **Wave 4** | | **Wave 5** | | **Wave 6** | |
|---|---|---|---|---|---|---|---|---|---|---|---|---|
| | Slow N (%) | Fast N (%) | Slow N (%) | Fast N (%) | Slow N (%) | Fast N (%) | Slow N (%) | Fast N (%) | Slow N (%) | Fast N (%) | Slow N (%) | Fast N (%) |
| **White UK** | 3,007 (40.27) | 4,460 (59.73) | 1,294 (38.22) | 2,092 (61.78) | 1,096 (38.70) | 1,736 (61.30) | 803 (41.18) | 1,147 (58.82) | 1,161 (42.37) | 1,579 (57.63) | 2,091 (42.94) | 2,779 (57.06) |
| **White overseas** | 459 (43.30) | 601 (56.70) | 180 (40.09) | 269 (59.91) | 151 (39.84) | 228 (60.16) | 86 (37.89) | 141 (62.11) | 120 (39.22) | 186 (60.78) | 346 (36.77) | 595 (63.23) |
| **Asian UK** | 375 (43.81) | 481 (56.19) | 154 (40.85) | 223 (59.15) | 121 (39.93) | 182 (60.07) | 79 (40.03) | 114 (59.07) | 90 (39.82) | 136 (60.18) | 235 (44.93) | 288 (55.07) |
| **Asian overseas** | 845 (58.28) | 605 (41.72) | 281 (54.88) | 231 (45.12) | 210 (52.90) | 187 (47.10) | 127 (55.95) | 100 (44.05) | 185 (54.90) | 152 (45.10) | 755 (56.64) | 578 (43.36) |

| | | | | | | | | | | | | |
|---|---|---|---|---|---|---|---|---|---|---|---|---|
| **Black UK** | 76 (50.67) | 74 (49.33) | 28 (51.85) | 26 (48.15) | 23 (43.40) | 30 (56.60) | 17 (50.00) | 17 (50.00) | 29 (50.88) | 28 (49.12) | 80 (53.69) | 69 (46.31) |
| **Black Overseas** | 215 (58.90) | 150 (41.10) | 60 (52.63) | 54 (47.37) | 54 (58.70) | 38 (41.30) | 23 (48.94) | 24 (51.06) | 45 (59.21) | 31 (40.79) | 405 (59.12) | 280 (40.88) |
| **Mixed UK** | 147 (38.99) | 230 (61.01) | 60 (38.96) | 94 (61.04) | 54 (39.42) | 83 (60.58) | 40 (43.48) | 52 (56.52) | 46 (36.22) | 81 (63.78) | 87 (35.66) | 157 (64.34) |
| **Mixed Overseas** | 67 (50.76) | 65 (49.24) | 30 (57.69) | 22 (42.31) | 22 (52.38) | 20 (47.62) | 11 (61.11) | 7 (38.89) | 21 (51.22) | 20 (48.78) | 52 (45.22) | 63 (54.78) |
| **Other UK** | 23 (44.23) | 29 (55.77) | 7 (26.92) | 19 (73.08) | 6 (35.29) | 11 (64.71) | 1 (12.50) | 7 (87.50) | 6 (46.15) | 7 (53.85) | 15 (31.25) | 33 (68.75) |
| **Other Overseas** | 111 (60.33) | 73 (39.67) | 43 (55.13) | 35 (44.87) | 35 (63.64) | 20 (36.36) | 19 (59.38) | 13 (40.62) | 27 (62.79) | 16 (37.21) | 89 (51.74) | 83 (48.26) |
| **Total** | 5,325 (44.03) | 6,768 (55.97) | 2,137 (41.08) | 3,065 (58.92) | 1,772 (41.14) | 2,535 (58.86) | 1,206 (42.64) | 1,622 (57.36) | 1,730 (43.62) | 2,236 (56.38) | 4,155 (45.76) | 4,925 (54.24) |
| **Medical Staff** | 1,195 (42.00) | 1,650 (58.00) | 484 (37.29) | 814 (62.71) | 379 (36.06) | 672 (63.94) | 254 (37.03) | 432 (62.97) | 325 (35.71) | 585 (64.29) | 580 (38.46) | 928 (61.54) |
| **Nurses, NAs, Midwives** | 1,259 (49.26) | 1,297 (50.74) | 530 (48.05) | 573 (51.95) | 457 (48.21) | 491 (51.79) | 305 (49.92) | 306 (50.08) | 483 (52.39) | 439 (47.61) | 1,262 (52.37) | 1,148 (47.63) |
| **AHPs (not including scientists)** | 1,546 (41.43) | 2,186 (58.57) | 579 (38.24) | 935 (61.76) | 496 (39.55) | 758 (60.45) | 344 (41.10) | 493 (58.90) | 493 (41.64) | 691 (58.36) | 1,071 (43.41) | 1,396 (56.59) |
| **Pharmacy** | 117 (47.95) | 127 (52.05) | 52 (52.53) | 47 (47.47) | 44 (49.44) | 45 (50.56) | 27 (50.94) | 26 (49.06) | 33 (47.83) | 36 (52.17) | 146 (40.90) | 211 (59.10) |
| **Healthcare Scientist** | 246 (44.48) | 307 (55.52) | 102 (41.13) | 146 (58.87) | 91 (40.81) | 132 (59.19) | 55 (41.35) | 78 (58.65) | 72 (43.90) | 92 (56.10) | 161 (43.28) | 211 (56.72) |
| **Ambulance** | 205 (46.28) | 238 (53.72) | 76 (43.68) | 98 (56.32) | 67 (42.14) | 92 (57.86) | 38 (40.86) | 55 (59.14) | 72 (48.98) | 75 (51.02) | 158 (46.88) | 179 (53.12) |
| **Dental** | 318 (41.95) | 440 (58.05) | 120 (39.47) | 184 (60.53) | 92 (39.32) | 142 (60.68) | 61 (39.61) | 93 (60.39) | 91 (42.72) | 122 (57.28) | 274 (45.36) | 330 (54.64) |
| **Optical** | 130 (43.77) | 167 (56.23) | 62 (43.97) | 79 (56.03) | 41 (42.71) | 55 (57.29) | 22 (41.51) | 31 (58.49) | 40 (45.98) | 47 (54.02) | 57 (42.86) | 76 (57.14) |

| | | | | | | | | | | | | |
|---|---|---|---|---|---|---|---|---|---|---|---|---|
| **Administrative** | 113 (46.89) | 128 (53.11) | 56 (42.42) | 76 (57.58) | 45 (47.37) | 50 (52.63) | 32 (43.24) | 42 (56.76) | 40 (44.44) | 50 (55.56) | 191 (52.91) | 170 (47.09) |
| **Other** | 196 (46.23) | 228 (53.77) | 76 (40.21) | 113 (59.79) | 60 (37.97) | 98 (62.03) | 68 (50.75) | 66 (49.25) | 81 (45.00) | 99 (55.00) | 255 (48.02) | 276 (51.98) |
| **Total** | 5,325 (44.03) | 6,768 (55.97) | 2,137 (41.08) | 3,065 (58.92) | 1,772 (41.14) | 2,535 (58.86) | 1,206 (42.64) | 1,622 (57.36) | 1,730 (43.62) | 2,236 (56.38) | 4,155 (45.76) | 4,925 (54.24) |

**Supplementary Table 6: Unadjusted Odds Ratios for total Physical Activity (GPPAQ) score**

| | Unadjusted model | | |
|---|---|---|---|
| | Unadjusted OR | 95% CI | *p*-value |
| White UK | REF | | |
| White born overseas | 1.26 | 1.17 - 1.37 | <0.001 |
| Asian born UK | 0.94 | 0.86 - 1.03 | 0.189 |
| Asian born overseas | 0.96 | 0.90 – 1.03 | 0.277 |
| Black Born UK | 0.80 | 0.66 – 0.96 | 0.016 |
| Black Born Overseas | 0.78 | 0.69 - 0.87 | <0.001 |
| Mixed Born UK | 1.08 | 0.95 - 1.23 | 0.213 |
| Mixed born Overseas | 0.91 | 0.74 - 1.12 | 0.356 |
| Other born UK | 1.19 | 0.86 - 1.64 | 0.300 |
| Other born Overseas | 0.78 | 0.63 - 0.88 | 0.001 |
| Medical staff | REF | | |
| Nurses, NAs, Midwives | 0.77 | 0.72 - 0.82 | <0.001 |
| AHPs (not including scientists) | 1.07 | 1.01 - 1.14 | 0.027 |
| Pharmacy | 0.78 | 0.68 - 0.90 | 0.001 |
| Healthcare Scientist | 0.67 | 0.60 - 0.74 | <0.001 |
| Ambulance | 1.99 | 1.72 - 2.28 | <0.001 |
| Dental | 0.84 | 0.76 - 0.93 | <0.001 |
| Optical | 0.83 | 0.71 – 0.97 | 0.020 |
| Administrative | 0.39 | 0.34 - 0.45 | <0.001 |
| Other | 0.62 | 0.55 - 0.69 | <0.001 |
| Male | REF | | |
| female | 0.70 | 0.67 – 0.74 | <0.001 |
| 20 - <30 years | REF | | |
| 30 - <40 years | 0.70 | 0.64 - 0.77 | <0.001 |
| 40 - <50 years | 0.57 | 0.51 - 0.62 | <0.001 |
| 50 - <60 years | 0.51 | 0.46 - 0.56 | <0.001 |
| >60 years | 0.48 | 0.44 - 0.53 | <0.001 |

| | | | |
|---|---|---|---|
| Q1 | REF | | |
| Q2 | 1.16 | 1.08 - 1.24 | <0.001 |
| Q3 | 1.17 | 1.09 - 1.26 | <0.001 |
| Q4 | 1.15 | 1.05 - 1.26 | 0.002 |
| Q5 | 1.26 | 1.17 - 1.36 | <0.001 |
| Q6 | 1.33 | 1.26 - 1.41 | <0.001 |

**Supplementary Table 7: Unadjusted Odds Ratios for total Exercise & Cycling only**

| | **Unadjusted model** | | |
|---|---|---|---|
| | Unadjusted OR | 95% CI | *p*-value |
| White UK | REF | | |
| White born overseas | 1.27 | 1.17 - 1.37 | <0.001 |
| Asian born UK | 0.96 | 0.88 - 1.05 | 0.428 |
| Asian born overseas | 0.83 | 0.77 - 0.87 | <0.001 |
| Black Born UK | 0.97 | 0.80 - 1.17 | 0.743 |
| Black Born Overseas | 0.54 | 0.48 - 0.61 | <0.001 |
| Mixed Born UK | 1.11 | 0.98 - 1.26 | 0.109 |
| Mixed born Overseas | 0.91 | 0.74 - 1.12 | 0.390 |
| Other born UK | 1.31 | 0.95 - 1.82 | 0.102 |
| Other born Overseas | 0.71 | 0.59 - 0.85 | <0.001 |
| Medical staff | REF | | |
| Nurses, NAs, Midwives | 0.52 | 0.47 – 0.57 | <0.001 |
| AHPs (not including scientists) | 0.90 | 0.85 – 0.96 | 0.001 |
| Pharmacy | 0.72 | 0.62 – 0.83 | <0.001 |
| Healthcare Scientist | 0.66 | 0.59 – 0.74 | <0.001 |
| Ambulance | 0.82 | 0.72 – 0.92 | 0.001 |
| Dental | 0.81 | 0.74 – 0.90 | <0.001 |
| Optical | 0.90 | 0.77 – 1.06 | 0.208 |
| Administrative | 0.52 | 0.45 – 0.60 | <0.001 |
| Other | 0.72 | 0.64 – 0.80 | <0.000 |
| Male | REF | | |

| | | | |
|---|---|---|---|
| female | 0.68 | 0.65 – 0.72 | <0.001 |
| 20 - <30 years | REF | | |
| 30 - <40 years | 0.78 | 0.71 – 0.85 | <0.001 |
| 40 - <50 years | 0.70 | 0.64 – 0.77 | <0.001 |
| 50 - <60 years | 0.71 | 0.65 – 0.78 | <0.001 |
| >60 years | 0.69 | 0.63 – 0.76 | <0.001 |
| Q1 | REF | | |
| Q2 | 1.33 | 1.25 – 1.42 | <0.001 |
| Q3 | 1.30 | 1.18 – 1.44 | <0.001 |
| Q4 | 1.33 | 1.17 – 1.50 | <0.001 |
| Q5 | 1.47 | 1.36 – 1.58 | <0.001 |
| Q6 | 1.28 | 1.21 – 1.35 | <0.001 |

**Supplementary Table 8: Unadjusted Odds Ratios for Walking Pace**

| | **Unadjusted model** | | |
|---|---|---|---|
| | Unadjusted OR | 95% CI | *p*-value |
| White UK | REF | | |
| White overseas | 1.03 | 0.96 - 1.11 | 0.410 |
| Asian UK | 0.93 | 0.85 – 1.01 | 0.07 |
| Asian overseas | 0.52 | 0.49 - 0.56 | <0.001 |
| Black UK | 0.66 | 0.55 - 0.79 | <0.001 |
| Black Overseas | 0.49 | 0.44 - 0.55 | <0.001 |
| Mixed UK | 1.110 | 0.97 - 1.24 | 0.126 |
| Mixed Overseas | 0.67 | 0.55 - 0.81 | <0.001 |
| Other UK | 1.25 | 0.91 - 1.72 | 0.170 |
| Other Overseas | 0.541 | 0.43 - 0.60 | <0.001 |
| Medical staff | REF | | |
| Nurses, NAs, Midwives | 0.63 | 0.59 - 0.67 | <0.001 |
| AHPs (not including scientists) | 0.90 | 0.85 – 0.96 | 0.001 |
| Pharmacy | 0.74 | 0.65 - 0.85 | <0.001 |
| Healthcare Scientist | 0.84 | 0.76 – 0.94 | 0.001 |
| Ambulance | 0.76 | 0.67 - 0.85 | <0.001 |
| Dental | 0.87 | 0.79 – 0.95 | 0.003 |
| Optical | 0.82 | 0.79 – 0.95 | 0.007 |

| | | | |
|---|---|---|---|
| Administrative | 0.68 | 0.60 - 0.78 | <0.001 |
| Other | 0.76 | 0.68 - 0.84 | <0.001 |
| Male | REF | | |
| female | 0.80 | 0.76 – 0.84 | <0.001 |
| 20 - <30 years | REF | | |
| 30 - <40 years | 0.67 | 0.63 - 0.75 | <0.001 |
| 40 - <50 years | 0.61 | 0.56 - 0.67 | <0.001 |
| 50 - <60 years | 0.54 | 0.50 - 0.59 | <0.001 |
| >60 years | 0.45 | 0.41 - 0.50 | <0.001 |
| Q1 | REF | | |
| Q2 | 1.12 | 1.06 - 1.21 | <0.001 |
| Q3 | 1.13 | 1.05 - 1.21 | 0.001 |
| Q4 | 1.06 | 0.97 - 1.15 | 0.180 |
| Q5 | 1.02 | 0.95 - 1.09 | 0.649 |
| Q6 | 0.93 | 0.88 - 0.99 | 0.012 |

**Supplementary Figures 1, 2 and 3. Sensitivity analysis for full cohort vs participants with 2 or more observations only**

Supplementary Figure 1: General Practice Physical Activity Questionnaire: Total Score

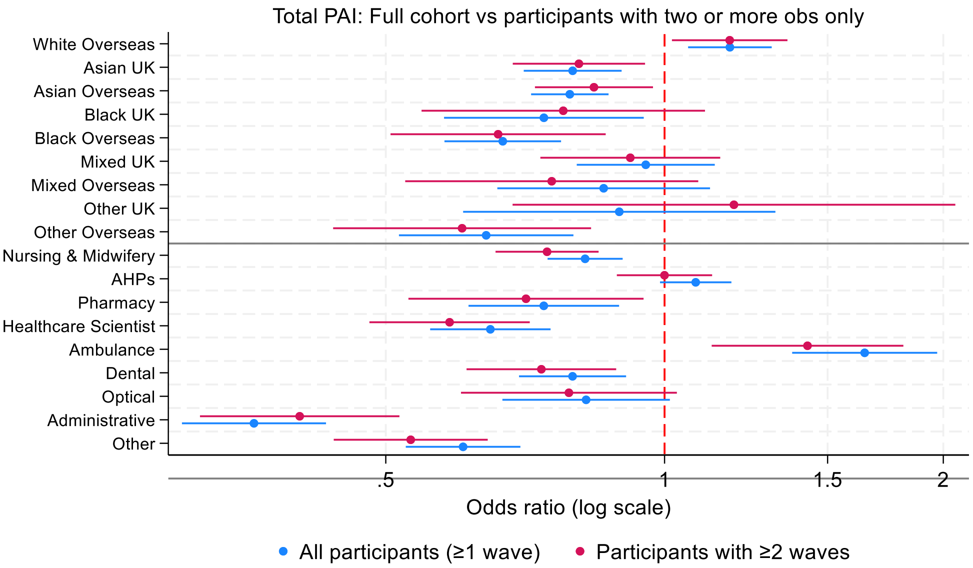

Supplementary Figure 2: General Practice Physical Activity Questionnaire: Exercise and cycling only

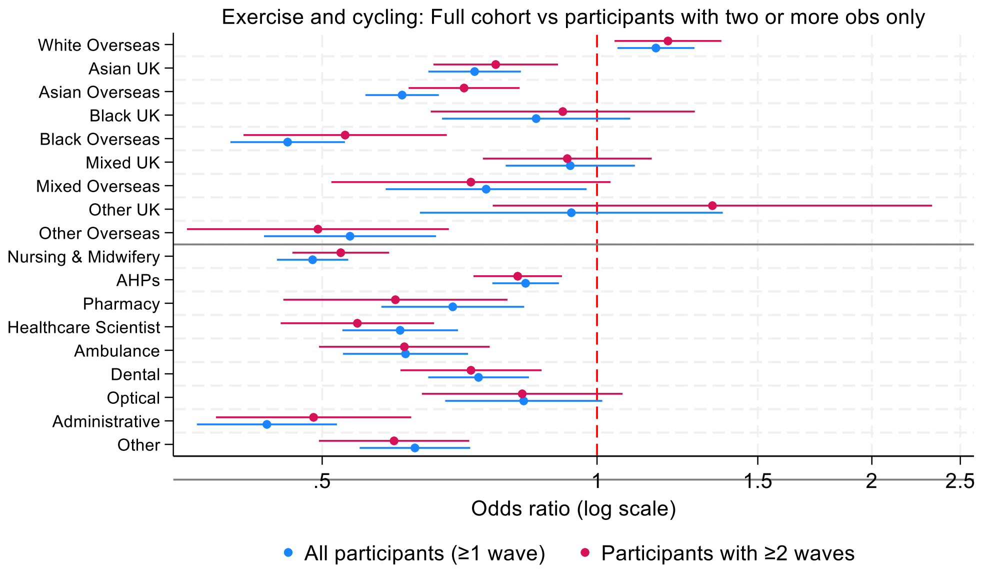

Supplementary Figure 3: General Practice Physical Activity Questionnaire: Walking Pace

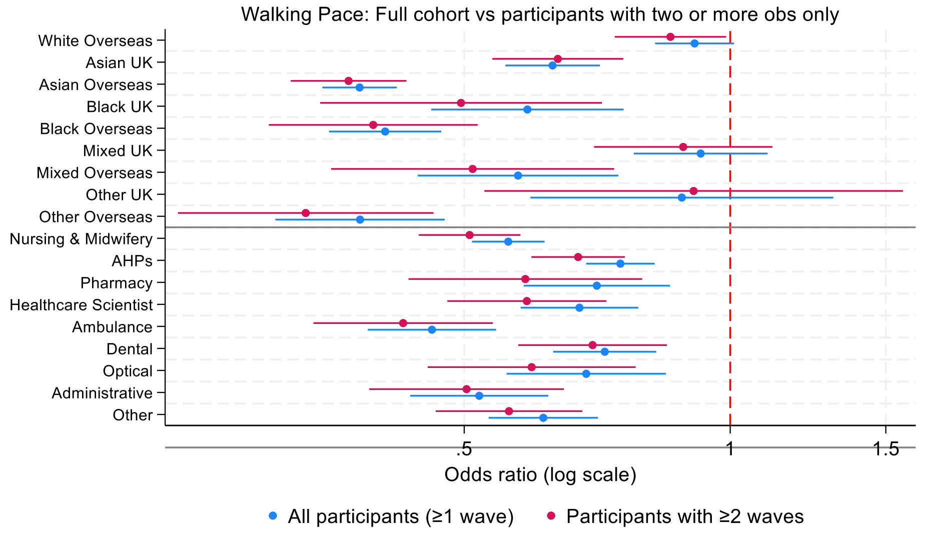
