## Supplementary material for "Demographic and occupational differences in physical activity and walking pace in UK healthcare workers: a nationwide longitudinal cohort study": Figure 3

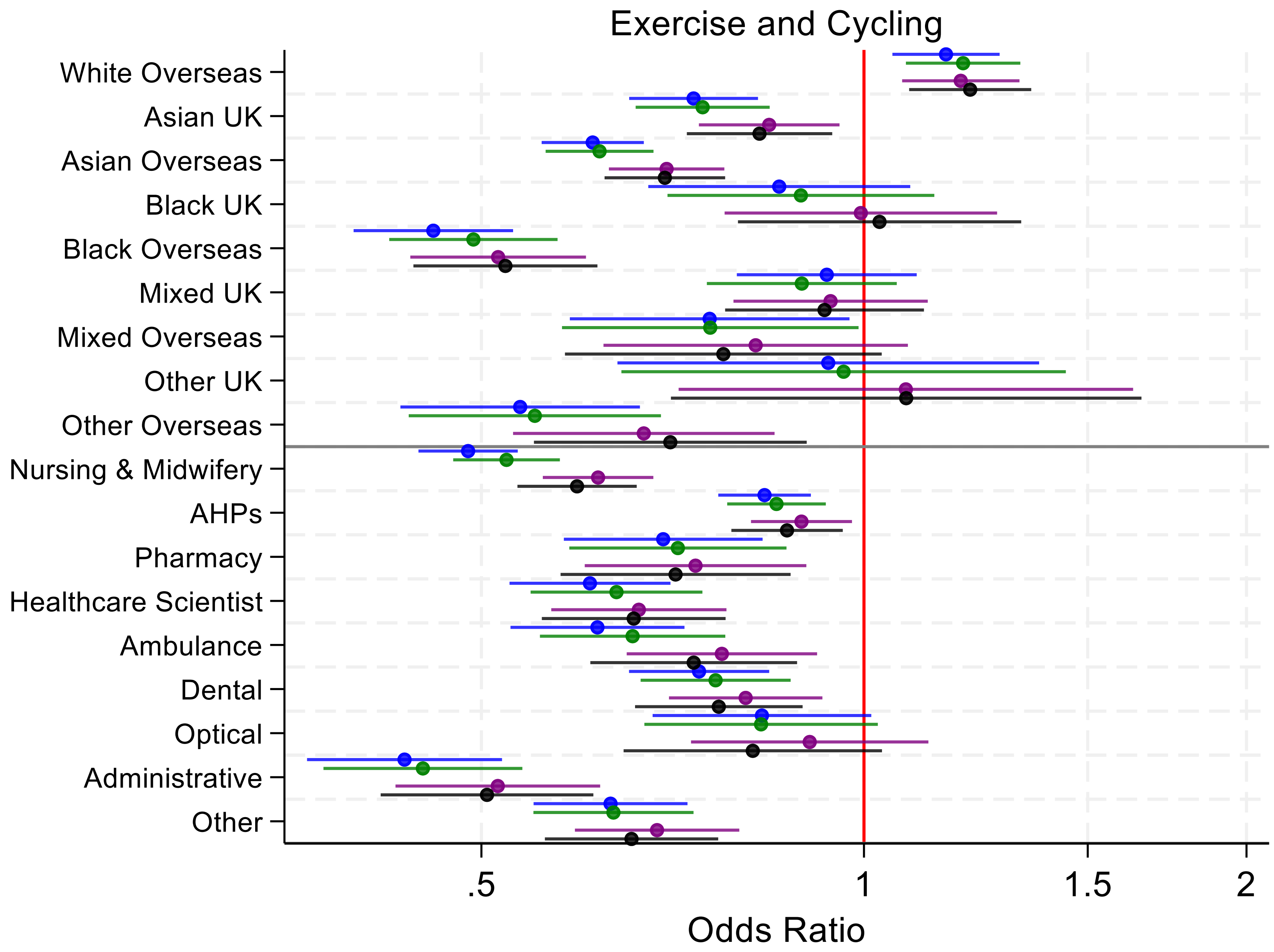


Model 1 = Ethnicity, job role, sex, age, wave.

Model 2 = Model 1 + IMD

Model 3 = Model 2 health/lifestyle outcomes (number of comorbidities, alcohol consumption, and smoking status).

Model 4 = Model 3 + shift variables (nights shifts and hours worked)
