## Supplementary figures and images for "Demographic and occupational differences in physical activity and walking pace in UK healthcare workers: a nationwide longitudinal cohort study"

### Figure 1

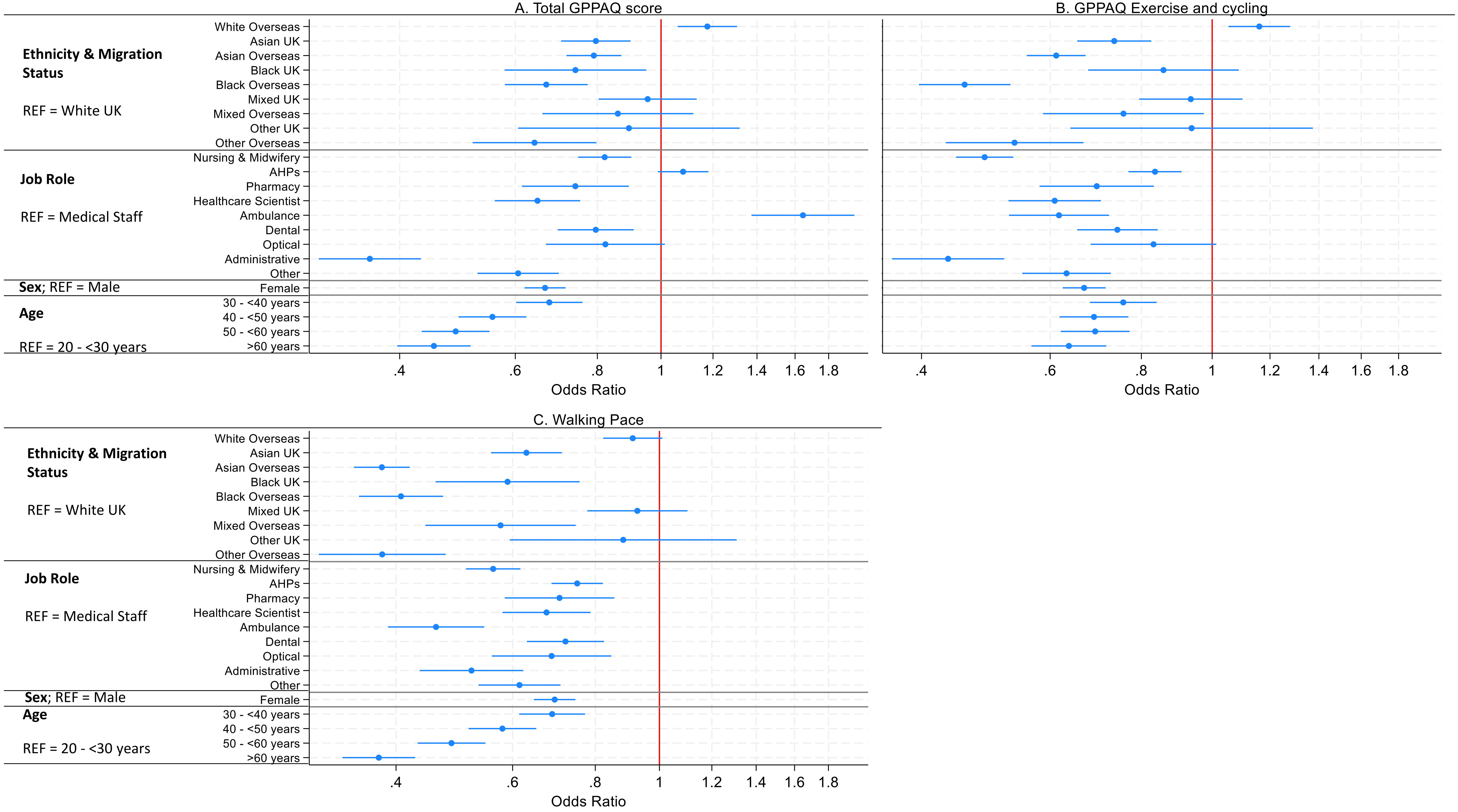

### Figure 2

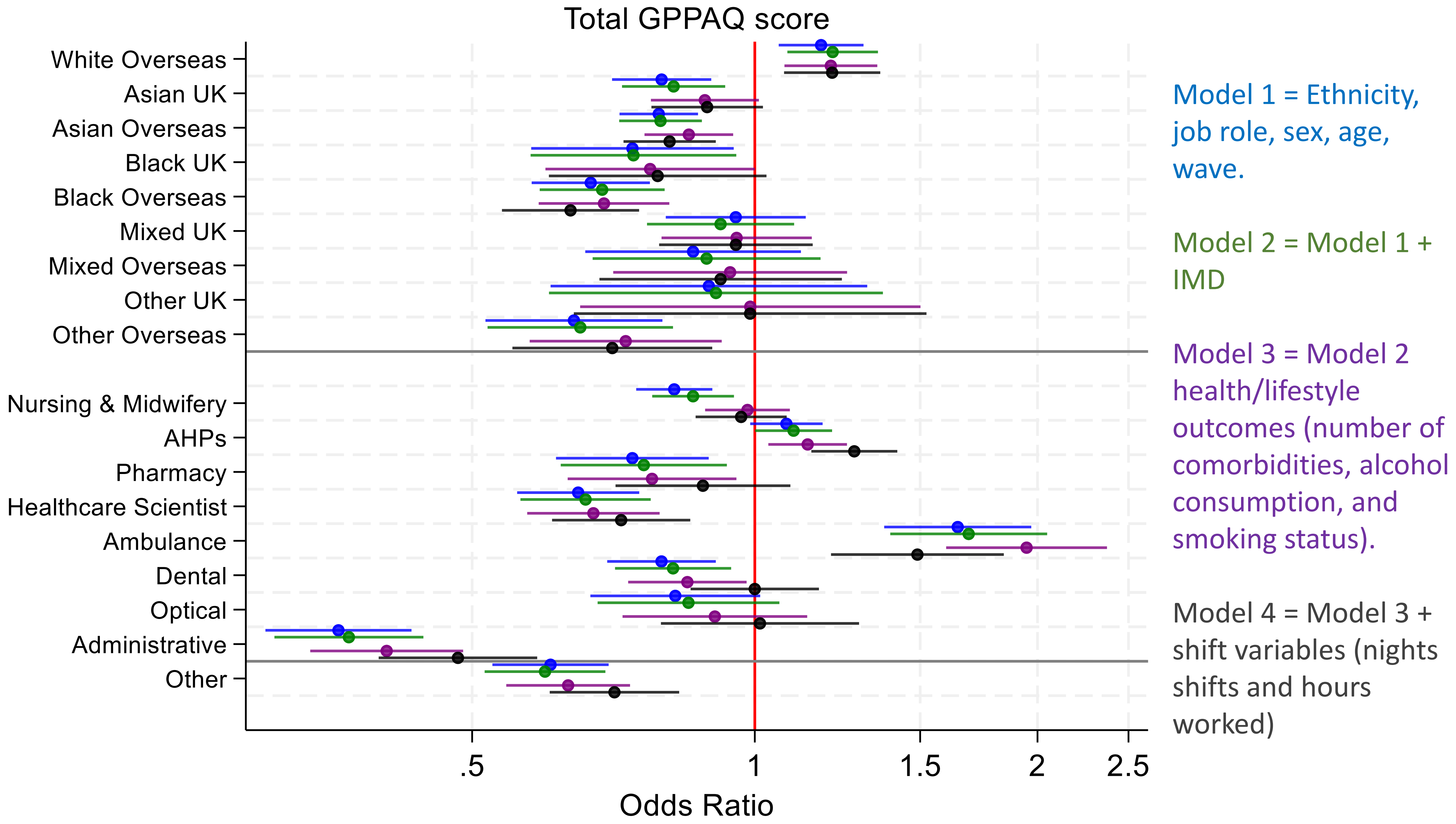
